# Construction and validation of a predictive model of mortality of tuberculosis-destroyed lung patients requiring mechanical ventilation: A single-center retrospective case-control study

**DOI:** 10.1101/2023.07.28.23293310

**Authors:** Kunping Cui, Yi Mao, Liangshuang Jiang, Yongli Zheng, Lang Yang, Yixiang Yang, Guihui Wu, Shenjie Tang

**Affiliations:** Intensive care unit, Public Health Clinical Center of Chengdu, Sichuan, China; Public Health Clinical Center of Chengdu, Sichuan, China; Tuberculosis department,Public Health Clinical Center of Chengdu, Sichuan, China; Tuberculosis department, Beijing Chest Hostpital capital University, Beijing, China

**Keywords:** tuberculosis-destroyed lung, mechanical ventilation;risk prediction model

## Abstract

**Background:** The mortality rate for intensive care unit (ICU) tuberculosis-destroyed lung (TDL) patients requiring mechanical ventilation remains high.

**Methods:** We conducted a retrospective analysis of adult TDL patients requiring mechanical ventilation who were admitted to the ICU of a tertiary infectious disease hospital in Chengdu, Sichuan Province, China from January 2019 to March 2023. Univariate and multivariate COX regression analyses were conducted to determine independent patient prognostic risk factors that were used to construct a predictive model of patient mortality.

**Results:** For the 331 study subjects, the median age was 63.0 (50.0-71.0) years, 262 (79.2%) were males and the mortality rate was 48.64% (161/331). Training and validation data sets were obtained from 245 and 86 patients, respectively. Analysis of the training data set revealed that a body mass index (BMI) of<18.5kg/m^2^, blood urea nitrogen level of ≥7.14mmol/L and septic shock were independent risk factors for increased mortality of TDL patients requiring mechanical ventilation. These variables were then used to construct a risk-based model for predicting patient mortality. Area under the curve,sensitivity and specificity values obtained using the model for the training data set were 0.808, 79.17% and 68.80%, respectively, and corresponding values obtained using the validation data set were 0.876, 95.12% and 62.22%. respectively. Results of concurrent correction curve and decision curve analyses verified the model possessed high predictiveability.

**Conclusions:** This model may facilitate early identification and classification-based clinical management of TDL patients requiring mechanical ventilation who are at high risk of death.

---

Tuberculosis(TB) is a chronic infectious disease caused by *Mycobacterium tuberculosis*. In 2021, about 10.6 million new TB cases and approximately 1.5 million TB-related deaths, of which 90% were caused by pulmonary TB, were reported globally[1]. Importantly, pulmonary TB is associated with a high lung injury rate approaching 68% [2–4], with lung injury leading to development of TB-destroyed lung (TDL) disease in 1.30% of cases[5].TDL is a consequence of improper anti-TB treatment management during the early TB disease stage that causes marked unilateral or bilateral erosion and/or deterioration of lung tissue structure that results in insufficient gas exchange area. As the disease progresses, lung function continues to decline until the normal exchange of inhaled air or oxygen is no longer adequate, resulting in irreversible pulmonary ventilation and ventilation dysfunction [6] that are often accompanied by different degrees of hypoxia, repeated secondary infections, hemoptysis, empyema and pneumothorax.Notably, the TDL patient mortality rate has been reported to be 28%[7–11], while the mortality rate of TDL patients requiring mechanical ventilation (TDL-MV)is much higher (61%)[11]. Therefore, early identification of risk factors affecting TDL-MV patient prognosis and clinical decision-making with regard to implementation of active interventions are of great importance for improving TDL-MV patient prognosis.

At present, Acute Physiological and Chronic Health Assessment (APACHE II) and Sequential Organ Failure Assessment(SOFA) scores are often used to assess disease severity of ICU patients with TDL-MV. However, these scoring systems do not provide consistent and accurate mortality risk estimates for patients with specific diseases and thus are of limited value when used for this purpose. For example, although APACHE II scoring has been widely used for conducting clinical early prognostic assessments of patients with certain critical diseases,this method requires collection of numerous types of data and thus is complicated, difficult to implement and performs poorly when used within 24 h of ICU admission[12]. SOFA scoringis also of limited value, since this method is mainly used to assess critically ill patients with sepsis or organ dysfunction [13,14]. Meanwhile,it has been reported that APACHE II and SOFA scoring-based methods cannot be used to accurately predict TDL-MV patient mortality rates [15]. Nevertheless, in the face of relentless global TB epidemics, increased clinical awareness of TB pulmonary injury has stimulated research to enhance understanding of its impact on patient outcomes, including a small number of studies based on small numbers of patients exploring risk factors affecting TDL-MV patient prognosis[7,11]. In order to build on the results of those limited studies, here we identified clinical characteristics and independent risk factors related to ICU TDL-MV patient mortality and used them to construct a predictive model. The resulting model should help clinicians achieve early prognosis and improved interventional treatments of TDL-MV patients in order to improve treatment outcomes for this patient population.

## MATERIALS AND METHODS

### Study design

The study was conducted at the Public Health Clinical Center of Chengdu, a 1,500-bed tertiary hospital specializing in infectious diseases in Chengdu, a city In southwestern China. The Research Ethics Committee of the center approved the study design (approval number:YJ-K2022-02-01) and exempted the study from the informed consent requirement. In this study, 499 prospective TDL patients seeking care at the above mentioned center from January 2019 to March 2023 were screened based on information obtained from the electronic information management system of Chengdu Public Health Clinical Medical Center. Thereafter, study subjects were selected based on study inclusion and exclusion criteria, resulting in enrollment of a total of 331 adult ICU patients with TDL-MV in the study. Of these patients, 245 who were admitted to the ICU between January 2019 and March 2022 were assigned to the model training group and the remaining 86 patients who were admitted to the ICU between April 2022 and March 2023 were assigned to the model validation group. A flow chart outlining the design of the study is presented in Figure 1. All TDL-MV patients enrolled in the study were diagnosed independently by at least three ICU medical specialists as based on patient epidemiological history, clinical manifestations and auxiliary examination findings (e.g., laboratory, imaging, pathological test results, etc.).TDL patients were diagnosed based onstandard clinical TDL diagnostic criteria as indicated in the guidance document WS288-2017 *Tuberculosis Diagnosis published in China in 2017*, which include the following: chest imaging examinations showing reduced lung tissue volume, secondary bronchiectasis or multiple fibrous thick-wall cavities and calcification, thoracic collapse, traction and displacement of adjacent hilum and mediastinum structures, pleural thickening and adhesion, etc.

**FIGURE 1.**
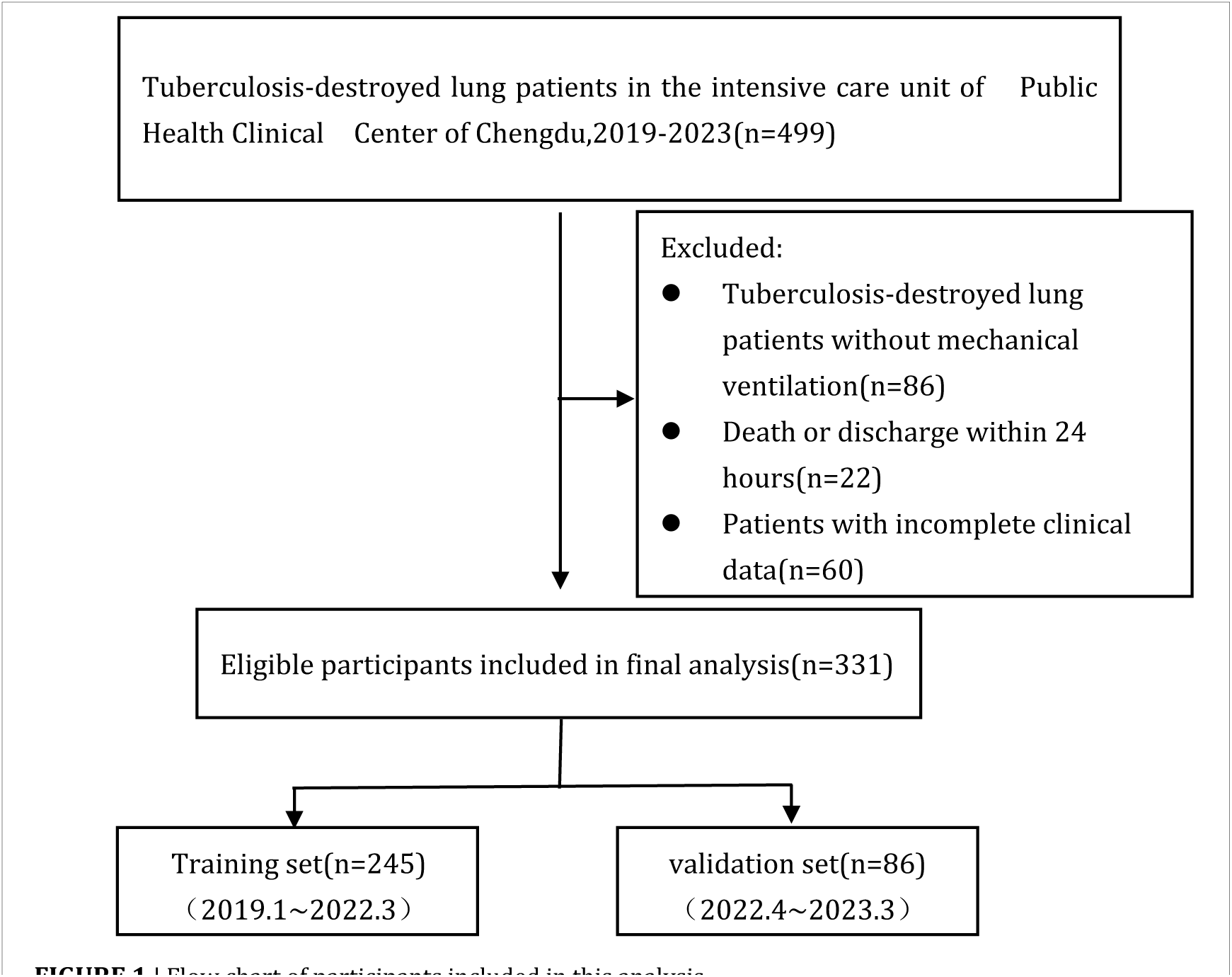
Flow chart of participants included in this analysis

MV therapeutic indications: (1)Loss of consciousness due to acute respiratory failure; (2) Respiratory rate >35∼40 times/min or <6∼8 breaths/min, abnormal respiratory rhythm or weak or absent spontaneous respiration; (3)PaO_2_< 50mmHg(especially after inhalation of oxygen); (4) Progressive increase of arterial PCO_2_level and pH decrease; (5) Respiratory failure after conventional treatmentand/or a trend of worsening respiratory function.

Inclusion criteria: (1)Patients ≥18 years of age, (2)Patients meeting both TDL diagnostic criteria and MV therapeutic indications.

Exclusion criteria: (1)Younger than 18 years of age;(1)TDL patients who did not require MV; (3) Patients with end-stage disease, irreversible disease and patients who could not benefit from ICU treatment; (4) Patients who died or were discharged within 24 h; (5) Patients with incomplete clinical data.

### Clinical data and laboratory test results

To determine risk factors that are associated with TDL-MV patient mortality, the following data were collected: (1)Patient demographic characteristics, including gender,age,race,residence,BMI,marriage history, smoking and alcohol consumption history. (2) TB epidemiological characteristics, including bacillus Calmette-Guérin (BCG) vaccination history and history of TB and/or drug-resistant TB infection. (3) Complications, including hemoptysis, pneumothorax and septic shock. (4) Comorbidities, including hypertension, diabetes, chronic liver disease, chronic kidney disease, HIV positive and chronic pulmonary heart disease. (5) Critical illness score obtained within 24 h after check-in based on APACHE II or SOFA scoring criteria. (6) Laboratory examination findings obtained within 24 h after check-in that included blood levels of white blood cells, red blood cells,hemoglobin,platelets, alanine aminotransferase, aspartate aminotransferase, total bilirubin, albumin, blood urea nitrogen (BUN), serum creatinine, pH, arterial partial pressure of carbon dioxide(PCO_2_), ratio of partial pressure of arterial oxygen to fraction of inspired oxygen(PO_2_/FiO_2_),lactic acid and levels of CD3+ T cells, CD4+ T cells and CD8+ T cells. (7) Findings indicating damaged lung. (8) 30-day prognosis (survival versus non-survival).

### Construction and validation of prediction model

Patients who provided model training set data were assigned to survival and non-survival groups according to 30-day prognosis results. In order to identify variables with statistical and clinical significance, patient data were screened via univariate analysis (P < 0.05). Next,potential risk factors affecting TDL-MV patient prognosis were identified using univariate COX regression analysis (P < 0.1)then independent prognostic risk factors were screened using stepwise multivariate COX regression analysis (P < 0.05).Thereafter, the prognostic model of TDL-MV patient mortality was generated based on the abovementioned mortality-related independent risk factors and their associated β coefficients.

The effectiveness of the model was internally verified using the training data set via three types of analysis: receiver operating characteristic (ROC) curve analysis (prediction model, APACHE II score and SOFA score),calibration curve analysis and decision curve analysis. Next, the validation data set was used to validate the model using the above mentioned analyses then the clinical efficacy of the model for predicting patient mortality was evaluated.Model-based predictions related to TDL-MV patient mortality were next stratified into low-risk and high-risk predictions of mortality according to scores obtained using the training set then Kaplan-Meier survival curves were plotted based on mortality rates obtained from training and validation data sets.

### Statistical analysis

Data were analyzed using IBM SPSS Statistics 23.0 statistical data analysis software (IBM, Armonk, New York, USA). ROC curve and survival curve analyses were conducted using MedCalc (MedCalc Software, Ltd., Ostend, Belgium). Calibration curves and decision curves were drawn using R software (version 4.5.3,https://CRAN.R-project.org, R Foundation, Vienna, Austria).the calibration curves was plotted used the “rms”package.the decision curve analysis was performed with the “rmda” package.

The Kolmogorov-Smirnov test was used for normality testing of continuous variables then normally distributed continuous variables were expressed as the mean ± standard deviation (SD). The independent sample t test was used to conduct comparisons between groups.

Non-normally distributed continuous variables were expressed as quartile median values and inter quartile range (IQR) values. Comparisons between groups were conducted using the Mann-Whitney U test. Categorical variables were analyzed using the Chi-square test for continuity correction (χ^2^). For univariate Cox regression analysis, variables with P values of<0.1 were selected for multivariate Cox regression analysis; those with P values of <0.05 were considered statistically significant. Survival curves were plotted using the Kaplan-Meier method.

### Demographic and clinical characteristics of the study population

Ultimately, 331 TDL-MV patients who met all inclusion and exclusion criteria were included in the study, with demographic and clinical characteristics of all subjects shown in Table 1. The overall median age of subjects was 63.0(50.0-71.1) years. Study subjects included 262 males (79.2%) and 69 females (20.8%);269 subjects were members of the Han nationality (81.3%) and 214 were members of other nationalities (64.7%); 255 were married (77.0%); 203 were smokers (61.3%); 127 consumed alcoholic beverages (38.4%);82 (24.8%) previously received the BCG vaccine. Clinically,279 (84.3%) patients were diagnosed with TB based on positive clinical findings, 71 (21.5%) with drug-resistant TB, 22 (6.6%) with hemoptysis, 29 (8.8%) with pneumothorax, 107 (32.3%) with chronic pulmonary heart disease, 78 (23.6%) with septic shock and 161 (48.64%) died.

**TABLE 1.**
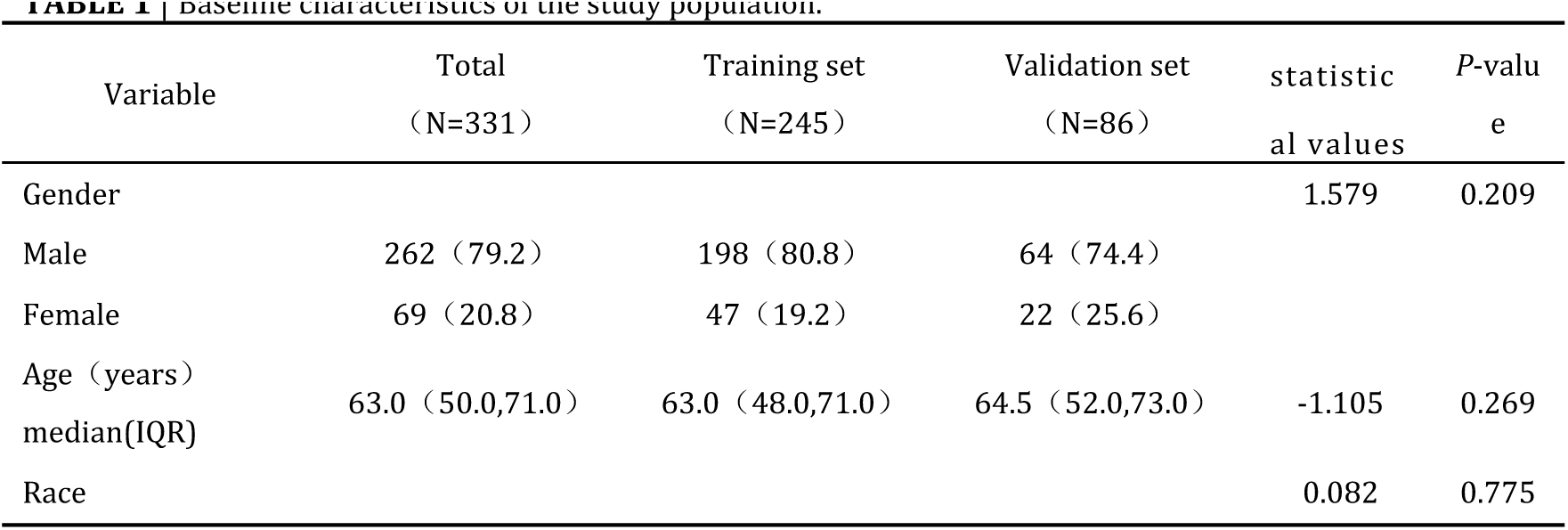
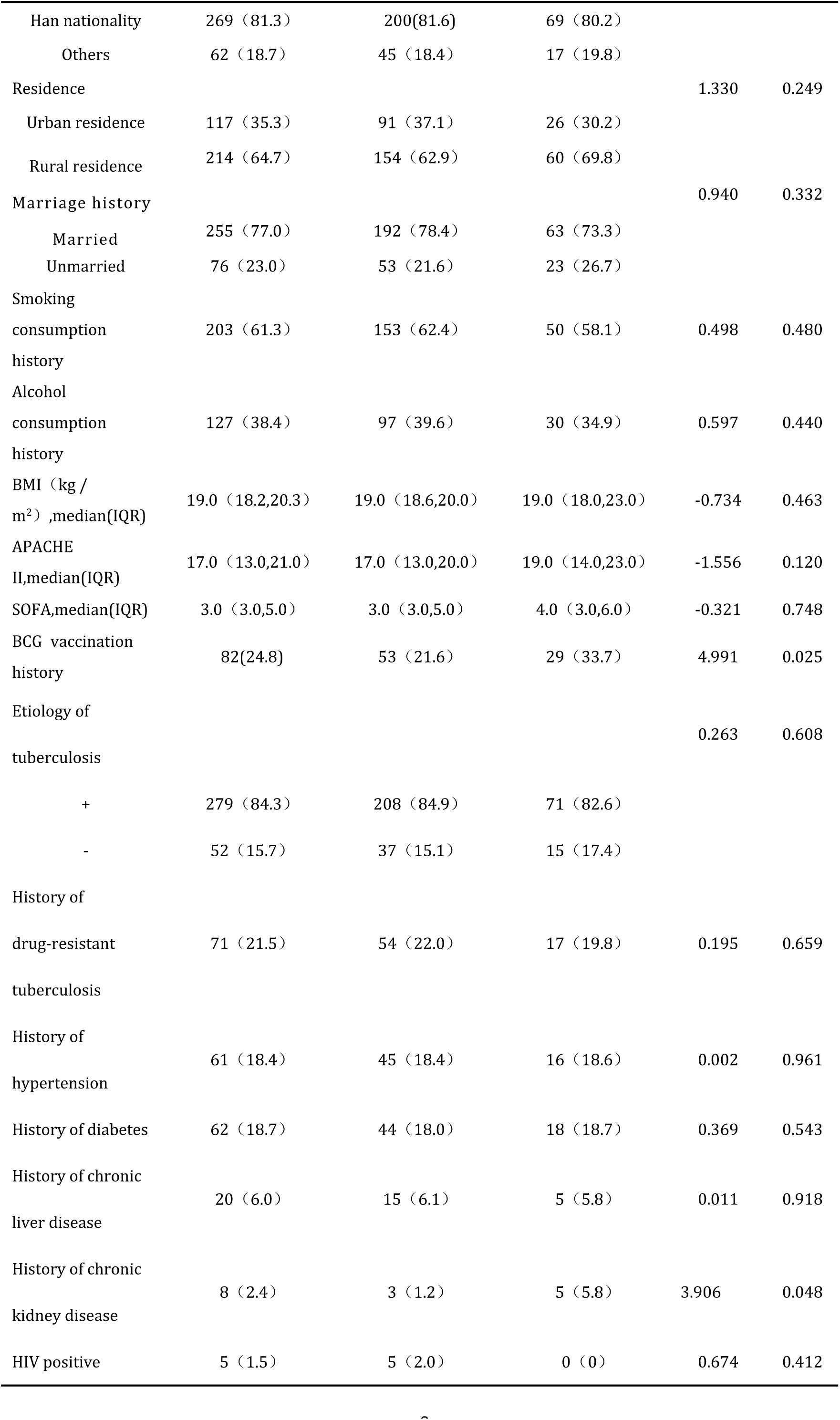
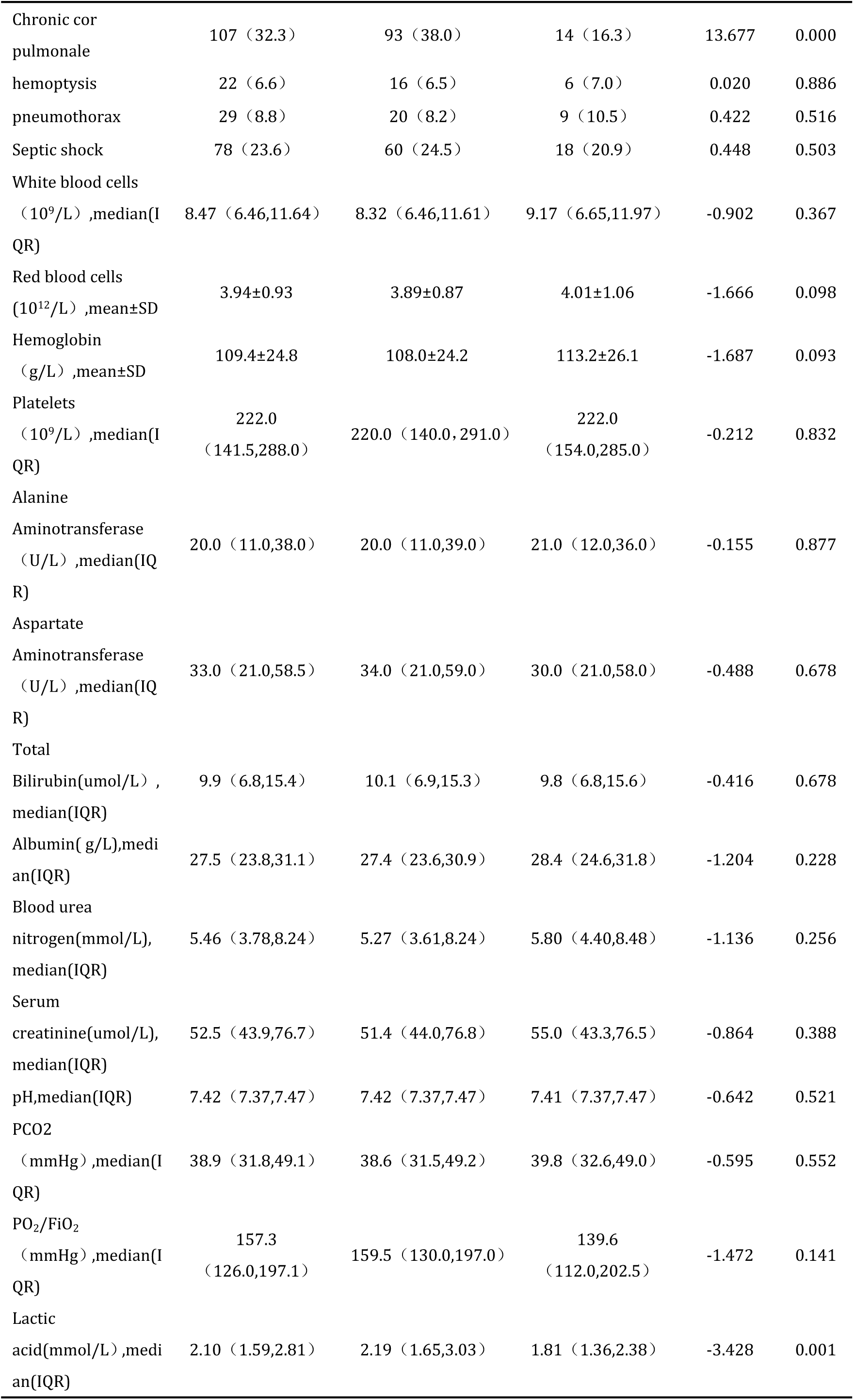
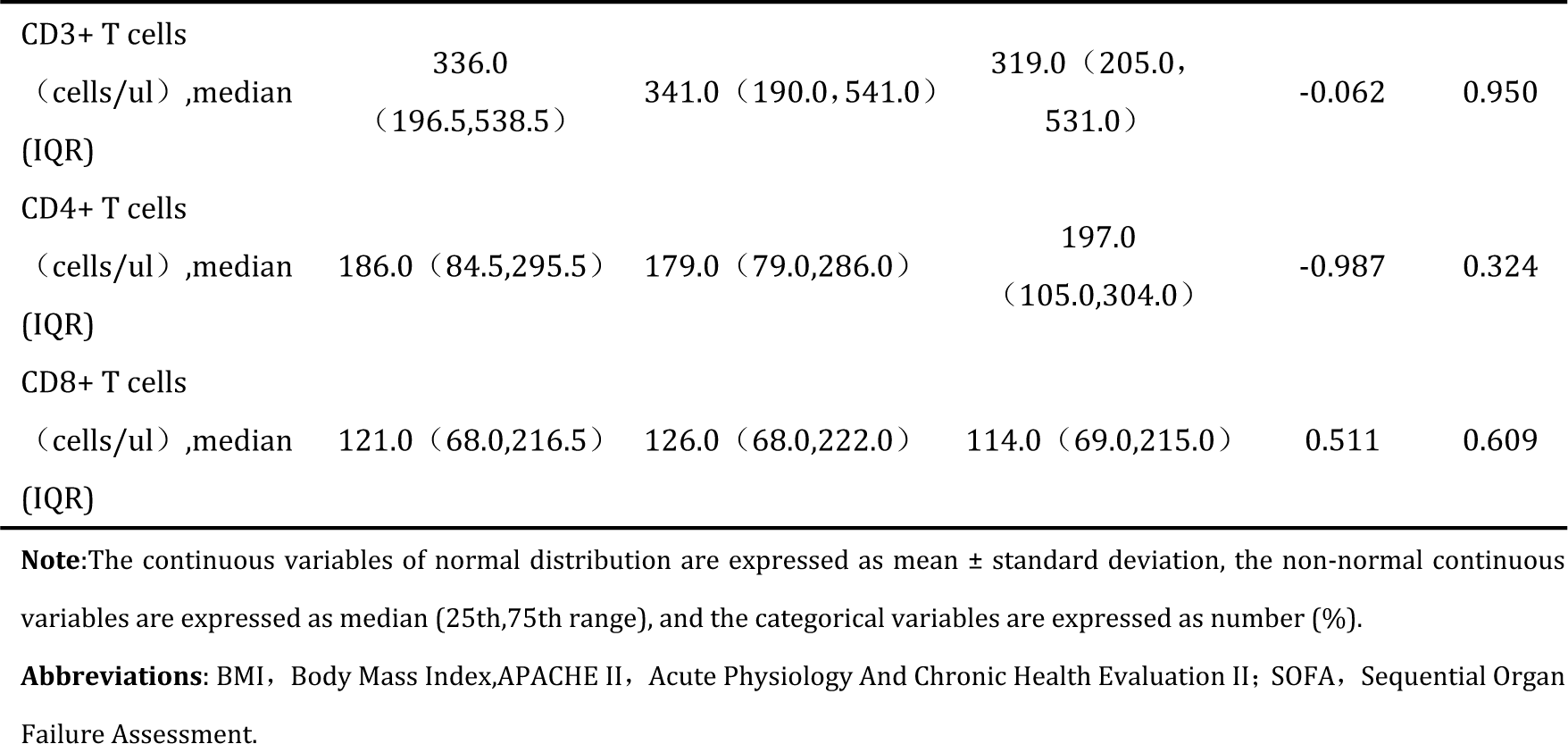
Baseline characteristics of the study population.

Of the 331 TDL-MVpatients, 28 were afflicted with miliary pulmonary TB with extrapulmonary TB involving the central nervous system TB (n=13), tuberculous pericarditis (n=45), TB of the abdominal cavity(n=35) and urinary TB (n= 7).No statistical differences in baseline characteristics were observed between the training set and the validation set except for BCG vaccination history, chronic kidney disease history, chronic pulmonary heart disease and blood lactic acid level (more detailed information is presented in Table 1).

### Construction of the mortality prediction model

Univariate analysis of training set data revealed 13 variables with statistically significant associations with patient mortality rate, including patient age, BMI, septic shock, and blood levels of platelets, serum albumin, urea nitrogen, serum creatinine, lactic acid, arterial blood pH, PO_2_/FiO_2_ and blood levels of CD3+T cells, CD4+ T cells and CD8+T cells (P < 0.05) (Table 2). Of these variables, those significantly associated with greater mortality (P values of <0.1) were identified using univariate COX regression analysis(detailed information presented in Table 3),including BMI, septic shock,PO_2_/FiO_2_, levels of serum albumin, BUN and serum creatinine and blood levels of CD8+T cells. Ultimately, variables with P values of<0.5,including BMI, septic shock and BUN (Table 4), were considered independent risk factors for increased TDL-MV patient mortality as based on regression β values (Table 5). Variables with β values of<1 received 1 point and those with β values of>1 received 2 points;independent variable scores for each patient were added together to obtain model-based mortality risk prediction scores that the minimum was 0 and the maximum was 4 pointes.

**TABLE 2.** Baseline characteristics of the study population in the training set.

| Variable | Total<br>(N=245) | Survivors<br>(N=125) | Non-survivors<br>(N=120) | statistic<br>al values | P-value |
| --- | --- | --- | --- | --- | --- |
| Gender |  |  |  | 0.413 | 0.521 |
| Male | 198 (80.8) | 103 (82.4) | 95 (79.2) |  |  |
| Female | 47 (19.2) | 22 (17.6) | 25 (20.8) |  |  |
| Age (years)<br>median(IQR) | 63.0 (48.0,71.0) | 59.0 (46.0,69.0) | 64.0 (51.0,74.5) | -2.227 | 0.026 |
| Race |  |  |  | 0.454 | 0.501 |
| Han nationality | 200 (81.6) | 100 (80.0) | 100 (83.3) |  |  |
| Others | 45 (18.4) | 25 (20.0) | 20 (16.7) |  |  |
| residence |  |  |  | 1.372 | 0.241 |
| Urban residence | 91 (37.1) | 42 (33.6) | 49 (40.8) |  |  |
| Rural residence | 154 (62.9) | 83 (66.4) | 71 (59.2) |  |  |
| Marriage history |  |  |  | 0.000 | 0.990 |
| Married | 192 (78.4) | 98 (78.4) | 94 (78.3) |  |  |
| Unmarried | 53 (21.6) | 27 (21.6) | 26 (21.7) |  |  |
| Smoking consumption<br>history | 153 (62.3) | 77 (61.6) | 76 (63.3) | 0.078 | 0.779 |
| Alcohol consumption<br>history | 97 (39.6) | 52 (41.6) | 45 (37.5) | 0.430 | 0.512 |
| BMI (kg /<br>m<sup>2</sup>) ,median(IQR) | 19.0 (18.6,20.0) | 19.3 (18.8,20.0) | 18.8 (17.8,19.5) | -4.361 | 0.000 |
| APACHE<br>II,median(IQR) | 17.0 (13.0,20.0) | 15.0 (12.0,18.0) | 18.5 (15.0,24.0) | -5.318 | 0.000 |
| SOFA,median(IQR) | 3.0 (3.0,5.0) | 3.0 (3.0,4.0) | 4.0 (3.0,7.0) | -5.707 | 0.000 |
| BCG vaccination<br>history | 53 (21.6) | 30 (24.0) | 23 (19.2) | 0.844 | 0.358 |
| Etiology of<br>tuberculosis |  |  |  | 0.098 | 0.754 |
| + | 208 (84.9) | 107 (85.6) | 101 (84.2) |  |  |
| - | 37 (15.1) | 18 (14.4) | 19 (15.8) |  |  |
| History of<br>drug-resistant<br>tuberculosis | 54 (22.0) | 29 (23.2) | 25 (20.8) | 0.200 | 0.655 |
| History of<br>hypertension | 45 (18.4) | 22 (17.6) | 23 (19.2) | 0.100 | 0.752 |
| History of diabetes | 44 (18.0) | 17 (13.6) | 27 (22.5) | 3.291 | 0.070 |
| History of chronic | 15 (6.1) | 11 (8.8) | 4 (3.3) | 3.183 | 0.074 |
| liver disease |  |  |  |  |  |
| History of chronic | 3 (1.2) | 0 (0) | 3 (2.5) | 1.434 | 0.231 |
| kidney disease |  |  |  |  |  |
| HIV positive | 5 (2.0) | 4 (3.2) | 1 (0.8) | 0.736 | 0.391 |
| Chronic cor pulmonale | 93 (38.0) | 51 (40.8) | 42 (35.0) | 0.875 | 0.350 |
| hemoptysis | 16 (6.5) | 9 (7.2) | 7 (5.8) | 0.187 | 0.665 |
| pneumothorax | 20 (8.2) | 12 (9.6) | 8 (6.7) | 0.703 | 0.402 |
| Septic shock | 60 (24.5) | 6 (4.8) | 54 (45.0) | 53.504 | 0.000 |
| White blood cells<br>($10^9/L$),median(IQR) | 8.32 (6.46,11.61) | 8.40<br>(6.36,11.44) | 8.08 (6.51,11.62) | -0.307 | 0.759 |
| Red blood cells<br>($10^{12}/L$),median(IQR) | 3.85 (3.37,4.40) | 110.0 (95.0,124.0) | 104.0 (88.0,121.0) | -1.715 | 0.086 |
| Hemoglobin<br>(g/L),mean±SD | 108.0±24.2 | 110.8±23.6 | 105.1±24.6 | 1.844 | 0.066 |
| Platelets<br>($10^9/L$),median(IQR) | 220.0<br>(140.0,291.0) | 238.0 (162.0,307.0) | 199.5<br>(112.5,264.0) | -2.536 | 0.011 |
| Alanine<br>Aminotransferase<br>(U/L),median(IQR) | 20.0 (11.0,39.0) | 22.0 (12.0, 39.0) | 19.0 (10.0, 40.5) | -0.909 | 0.363 |
| Aspartate<br>Aminotransferase<br>(U/L),median(IQR) | 34.0 (21.0,59.0) | 35.0 (22.0,56.0) | 32.0 (21.0,59.5) | -0.290 | 0.772 |
| Total<br>Bilirubin($\mu\text{mol}/L$),median(IQR) | 10.1 (6.9,15.3) | 9.7 (7.1,15.0) | 10.3 (6.7,15.7) | -0.330 | 0.741 |
| Albumin(g/L),median(IQR) | 27.4 (23.6,30.9) | 28.10 (24.90,31.80) | 26.55<br>(22.25,29.95) | -3.072 | 0.002 |
| Blood urea<br>nitrogen($\text{mmol}/L$),median(IQR) | 5.27 (3.61,8.24) | 4.49 (3.32,6.40) | 6.91 (4.51,9.71) | -4.849 | 0.000 |
| Serum<br>creatinine($\mu\text{mol}/L$),median(IQR) | 51.4 (44.0,76.8) | 50.0 (43.0,64.0) | 56.8 (45.8,86.0) | -2.982 | 0.003 |
| pH,median(IQR) | 7.42 (7.37,7.47) | 7.44 (7.39,7.48) | 7.41 (7.35,7.46) | -2.184 | 0.029 |
| PCO2<br>(mmHg),median(IQR) | 38.6 (31.5,49.2) | 37.9 (31.5,47.3) | 39.2 (31.5,53.2) | -0.345 | 0.730 |
| PO<sub>2</sub>/FiO<sub>2</sub><br>(mmHg) ,median(IQ<br>R) | 159.5<br>(130.0,197.0) | 169.7 (142.0,200.3) | 146.2<br>(123.3,187.4) | -3.509 | 0.000 |
| Lactic<br>acid(mmol/L) ,median<br>(IQR) | 2.19 (1.65,3.03) | 2.06 (1.60,2.62) | 2.46 (1.70,3.39) | -2.591 | 0.010 |
| CD3+ T cells<br>(cells/ul) ,median(IQ<br>R) | 341.0<br>(190.0,541.0) | 382.0 (209.0,579.0) | 294.5<br>(144.0,467.5) | -2.766 | 0.006 |
| CD4+ T cells<br>(cells/ul) ,median(IQ<br>R) | 179.0<br>(79.0,286.0) | 201.0<br>(102.0,325.0) | 156.0 (64.5,262.5) | -2.209 | 0.027 |
| CD8+ T cells<br>(cells/ul) ,median(IQ<br>R) | 126.0<br>(68.0,222.0) | 153.0 (77.0,265.0) | 110.5 (55.0,191.5) | -2.557 | 0.011 |

**Note:**The continuous variables of normal distribution are expressed as mean ± standard deviation, the non-normal continuous variables are expressed as median (25th,75th range), and the categorical variables are expressed as number (%).
**Abbreviations:** BMI, Body Mass Index,APACHE II, Acute Physiology And Chronic Health Evaluation II; SOFA, Sequential Organ Failure Assessment.

**TABLE 3.**
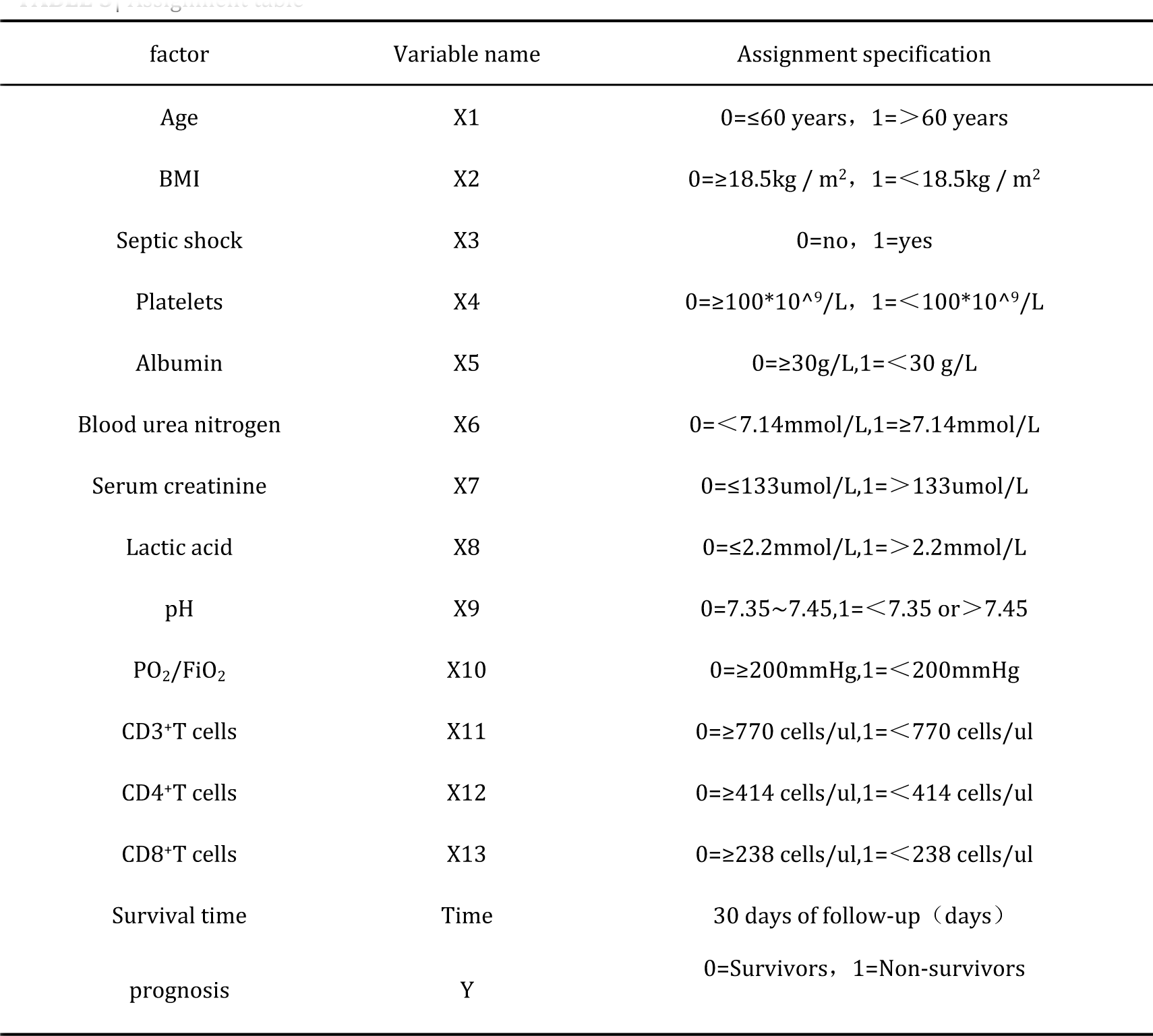
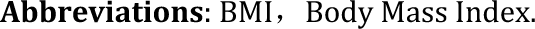
Assignment table.

| factor | Variable name | Assignment specification |
| --- | --- | --- |
| Age | X1 | 0=≤60 years, 1=$>$60 years |
| BMI | X2 | 0=≥18.5kg / m<sup>2</sup>, 1=$<$18.5kg / m<sup>2</sup> |
| Septic shock | X3 | 0=no, 1=yes |
| Platelets | X4 | 0=≥100*10<sup>9</sup>/L, 1=$<$100*10<sup>9</sup>/L |
| Albumin | X5 | 0=≥30g/L,1=$<$30 g/L |
| Blood urea nitrogen | X6 | 0=$<$7.14mmol/L,1=≥7.14mmol/L |
| Serum creatinine | X7 | 0=≤133umol/L,1=$>$133umol/L |
| Lactic acid | X8 | 0=≤2.2mmol/L,1=$>$2.2mmol/L |
| pH | X9 | 0=7.35~7.45,1=$<$7.35 or $>$7.45 |
| PO<sub>2</sub>/FiO<sub>2</sub> | X10 | 0=≥200mmHg,1=$<$200mmHg |
| CD3+T cells | X11 | 0=≥770 cells/ul,1=$<$770 cells/ul |
| CD4+T cells | X12 | 0=≥414 cells/ul,1=$<$414 cells/ul |
| CD8+T cells | X13 | 0=≥238 cells/ul,1=$<$238 cells/ul |
| Survival time | Time | 30 days of follow-up (days) |
| prognosis | Y | 0=Survivors, 1=Non-survivors |

**Abbreviations:** BMI, Body Mass Index.

**TABLE 4.** Univariate and multivariate Cox regression analysis of clinical factors in the training set.

| Variable | Univariate analysis |  | Multivariate analysis |  |  |
| --- | --- | --- | --- | --- | --- |
| | HR (95%CI) | P | $\beta$ | HR (95%CI) | P |
| Age | 1.295(0.900~1.861) | 0.163 |  |  |  |
| BMI | 2.931(2.013~4.266) | 0.000 | 0.762 | 2.144(1.447~3.176) | 0.000 |
| Septic shock | 4.491(3.105~6.497) | 0.000 | 1.226 | 3.407(2.309~5.028) | 0.000 |
| Platelets | 1.328(0.822~2.147) | 0.247 |  |  |  |
| Albumin | 1.577(1.043,2.384) | 0.031 |  |  |  |
| Blood urea nitrogen | 2.231(1.558~3.195) | 0.000 | 0.682 | 1.977(1.374~2.845) | 0.000 |
| Serum creatinine | 3.188(1.850~5.493) | 0.000 |  |  |  |
| Lactic acid | 1.338(0.934~1.916) | 0.112 |  |  |  |
| pH | 0.954(0.665~1.370) | 0.800 |  |  |  |
| PO<sub>2</sub>/FiO<sub>2</sub> | 1.531(0.964~2.432) | 0.071 |  |  |  |
| CD3<sup>+</sup>T cells | 1.471(0.718~3.015) | 0.292 |  |  |  |
| CD4<sup>+</sup>T cells | 1.623(0.850~3.102) | 0.143 |  |  |  |
| CD8<sup>+</sup>T cells | 1.615(1.008~2.587) | 0.046 |  |  |  |

**Abbreviations:** CI, confidence interval; HR, Harard ratio;BMI, Body Mass Index.

**TABLE 5.** Construction of a risk prediction model for death in patients with TDL-MV.

| Variable | score |
| --- | --- |
| BMI<18.5kg/m<sup>2</sup> | 1 |
| Septic shock | 2 |
| Blood urea nitrogen≥7.14mmol/L | 1 |
| Total score | 4 |

**Abbreviations:** BMI, Body Mass Index.

### Validation of the mortality prediction model

Using the training data set to test the risk prediction model revealed an area under the curve (AUC) value for 30-day TDL-MV patient mortality of 0.808, a sensitivity rate of 79.1% and a specificity rate of 68.80%. Meanwhile,the SOFA score AUC value was 0.702, the sensitivity rate was 49.17% and the specificity rate was 83.20%, an APACHE II score-based AUC value of 0.696, sensitivity rate of 50.00% and specificity rate of 77.60%(Figure 2A).

**FIGURE 2.**
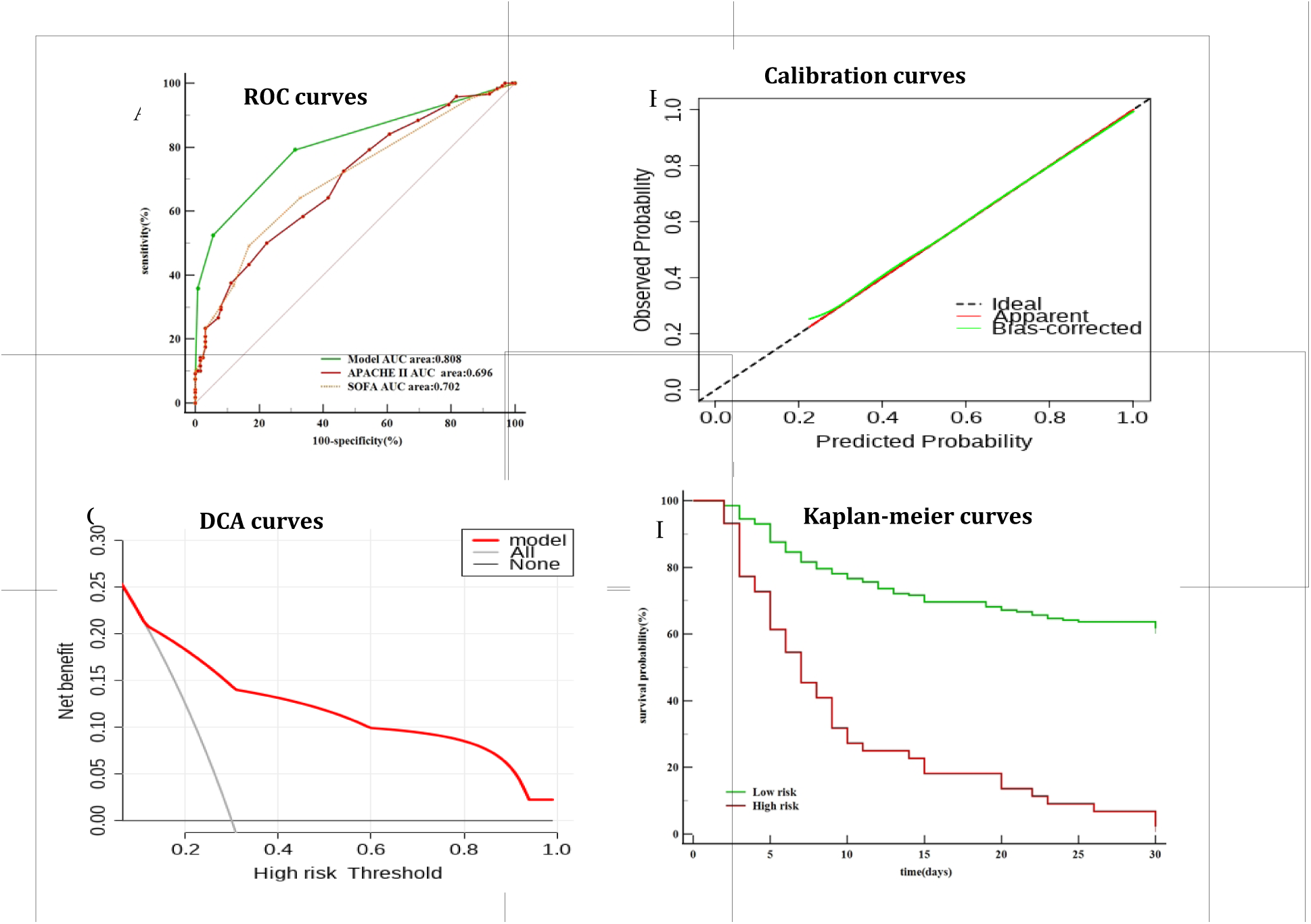
Tranining set |(A)Prediction model and other clinical scores ROC curve; (B)Calibration curves for Prediction model(B=100 repetitions,boot Mean absolute error=0.014 n=245).The horizontal axis was the predicted probability of 30-days death by the model, and the vertical axis was the actual probability. The dashed line indicates the predicted probability completely fits the actual probability;(C)Decision curve analysis for Prediction model. The y-axis measures the net benefit. The red line represents the Prediction model. The gray dotted line represents the assumption that all patients dead. The black dotted line represents the assumption that no patients dead.DCA:Decision curve analysis.(D)Kaplan-Meier survival curves of the patients with tuberculosis-destroyed lung requiring mechanical ventilation in training set. **Abbreviations**: APACHE II Acute Physiology And Chronic Health Evaluation II SOFA Sequential Organ Failure Assessment DCA Decision curve analysis.

Use of the validation data set to validate the mortality risk prediction model revealed an AUC value for 30-day TDL-MVpatient mortality of 0.876, a sensitivity rate of 95.12% and a specificity rate of 62.22%. Meanwhile, the SOFA score-based AUC value was 0.738, the sensitivity rate was 53.66% and the specificity rate was 84.44%, while the APACHE II score-based AUC value was 0.803, the sensitivity rate was 82.93% and the specificity rate was 66.67%(Figure 3A).

**FIGURE 3.**
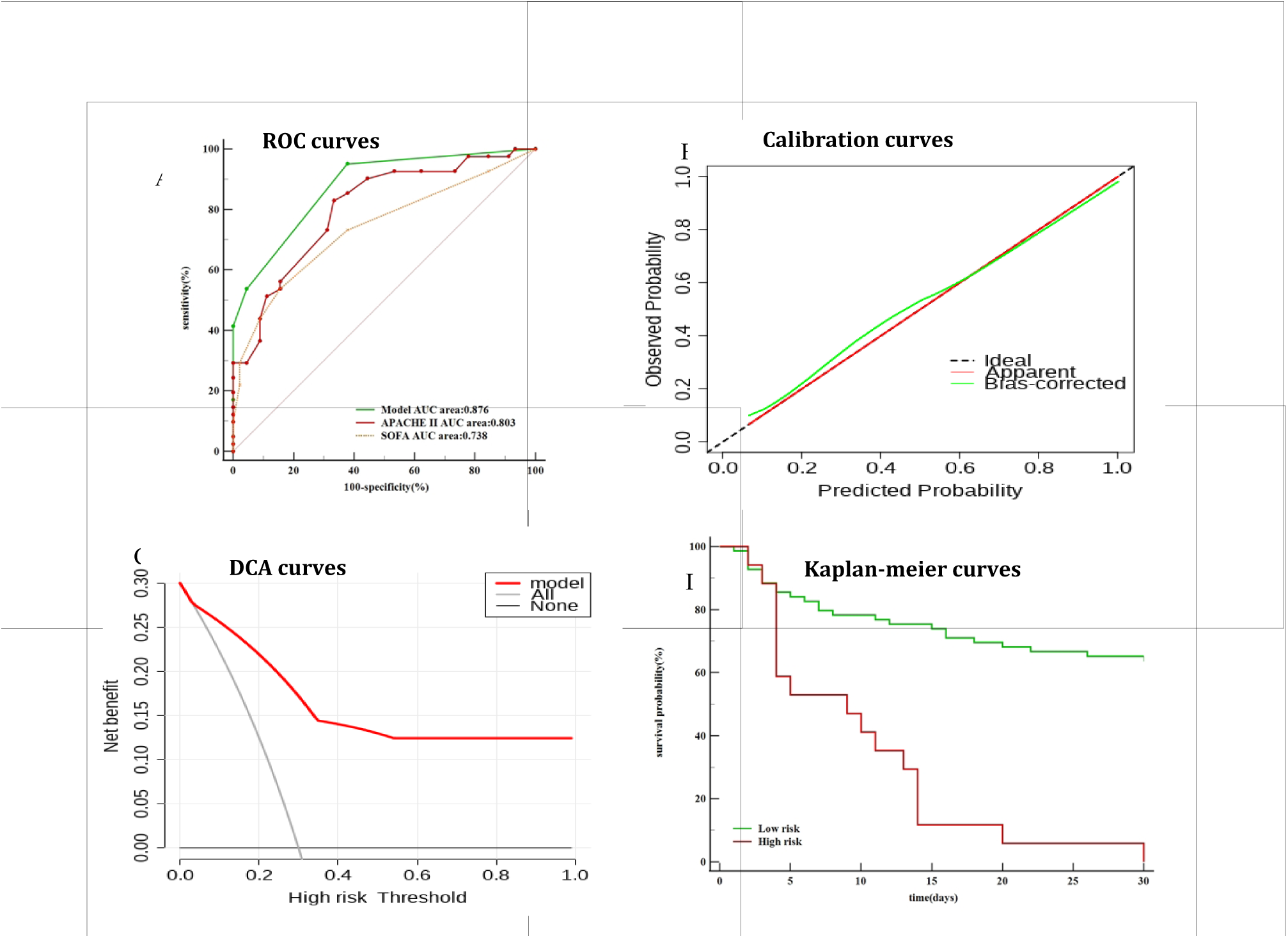
Validation set |(A)Prediction model and other clinical scores ROC curve; (B)Calibration curves for Prediction model(B=100 repetitions,boot Mean absolute error=0.023 n=86).The horizontal axis was the predicted probability of 30-days death by the model, and the vertical axis was the actual probability. The dashed line indicates the predicted probability completely fits the actual probability;(C)Decision curve analysis for Prediction model. The y-axis measures the net benefit. The red line represents the Prediction model. The gray dotted line represents the assumption that all patients dead. The black dotted line represents the assumption that no patients dead.DCA:Decision curve analysis.(D)Kaplan-Meier survival curves of the patients with tuberculosis-destroyed lung requiring mechanical ventilation in validation set. **Abbreviations**: APACHE II Acute Physiology And Chronic Health Evaluation II SOFA Sequential Organ Failure Assessment DCA Decision curve analysis.

Notably, predicted mortality rates obtained using the risk prediction model for both the training and validation data sets were close to the actual observed TDL-MV patient mortality rate, thus indicating good calibration curve agreement (Figure 2B, 3B). Moreover, results obtained via decision curve analysis also showed that the prediction model provided a good net benefit (Figure 2C, 3C).

### Prognostic comparison and risk stratification

Analysis of training data set-derived patient scores generated using the risk prediction model revealed an increase in TDL-MV patient mortality rate with increasing score. With regard to mortality risk, comparisons of patient scores to a critical threshold value resulted in the assignment of 44 patients to the high-risk group (3∼7 points) and 201 patients to the low-risk group (0∼2 points)(Table 6).An analysis of Kaplan-Meier curves revealed significant differences in survival between high-risk and low-risk patient groups asstratified using the risk prediction model developed usingboth training and validation data sets (Figure2D,3D).

**TABLE 6.** Mortality rates of the TDL-MV patients according to the risk prediction model. Trainingn set n=245 validation set(n=86)

| Score | Trainingn set (n=245) |  |  | validation set(n=86) |  |  |
| --- | --- | --- | --- | --- | --- | --- |
|  | Survivors<br>(N=125),<br>n | Non-survivors<br>(N=120),n | Mortality<br>(48.98%) | Survivors<br>(N=45),n | Non-survivors<br>(N=41),n | Mortality<br>(47.67%) |
| 0 | 86 | 25 | 22.5 | 28 | 2 | 6.6 |
| 1 | 32 | 32 | 50.0 | 15 | 17 | 53.1 |
| 2 | 6 | 20 | 76.9 | 2 | 5 | 71.4 |
| 3 | 1 | 34 | 97.1 | 0 | 10 | 100 |
| 4 | 0 | 4 | 100 | 0 | 7 | 100 |
| ≤2 | 124 | 77 | 38.3 | 45 | 24 | 34.7 |
| >2 | 1 | 43 | 97.7 | 0 | 17 | 100 |

## DISCUSSION

In this study, we constructed and validated a predictive model of mortality of tuberculosis-destroyed lung patients requiring mechanical ventilation.Internal verification of the risk prediction model indicated good discrimination and prediction performance of the model, thus demonstrating that use of the risk prediction model would not only enable more efficient utilization of limited medical resources, but would also reduce the TDL-MV patient mortality rate. Malnutrition has been shown in numerous studies to be associated with increased TB incidence and severity, anti-TB treatment failure, poorer TB treatment outcomes and increased mortality[16–19]. Results obtained in the current study showed that BMI, serum albumin level and blood CD4+ T cell level were significantly reduced in TDL-MV patients, with more pronounced reductions observed in the patient group with a high risk of death, BMI is an independent risk factor for TDL-MV patient death, which suggest that ineffective anti-TB treatment of patients with early-stage TB may cause long-term nutritional and immune depletionand significantly reduced anti-TB therapeutic responses after disease progression to late-stage TB disease,as consistent with results of previously reported studies of patients with severe lung destruction[20–22].Septic shock is likely induced capillary endothelial injury, tissue microcirculation ischemia and hypoxia. In turn,these effects can lead to vascular paralysis, organ function impairment and severe microcirculatory dysfunction.that can persist even after normalization of hemodynamics has been achieved through fluid resuscitation and administration of vasopressors and anti-infective therapies [23].which is one of several direct causes of ICU patient death and is associated with a mortality rate of about 50% [24–26]. The results of a retrospective nested cohort study reported in 2013 indicated that although clinical manifestations of M.tuberculosis infection-induced septic shock resembled those associated with other bacterial infections, the mortality rate of M.tuberculosis infection-induced septic shock(79.2%)was significantly greater than corresponding rates obtained for all other types of bacterial infections (49.7%)[27]. Indeed, in the current study the mortality rate of ICU TDL-MV patients with septic shock was markedly higher (∼92.3%), a result that may reflect the fact that our patients were late-stage TB patients and thus had more severe TB disease than patients in the above mentioned studies. BUN was used to assess renal impairment and protein catabolic function [28–31].A retrospective observational study reported in 2021 involving 845 patients with acute exacerbations of chronic obstructive pulmonary disease (AECOPD).which revealed that the elevated BUN level was associated with increased AECOPD patient mortality[32]. Meanwhile, results of an other retrospective study involving 2682 ICU patients with intracranial hemorrhage indicated that an elevated BUN level was significantly associated with poor prognosis[33]. In addition, results of other studies have confirmed that an elevated BUN level is associated with increased long-term mortality of severely ill patients[34]. Similarly, in our study of TDL-MV patients, the average BUN level of the patient group with greater risk of death exceeded that of the patient group with greater survival. Moreover, an increased BUN level was found to be an independent risk factor for increased TDL-MV patient mortality,The underlying mechanism needs further study.

In recent years, the mortality prediction of ICU patients has been widely investigated, but the general ICU severity predictive model were not sufficient for predicting mortality in the population of TDL-MV patients accurately and reliably. In our study,Analysis of risk prediction performance of the model using calibration curve and decision curves generated revealed that the model had excellent prediction ability and predicted patient mortality more accurately than did predictive methods based on traditional APACHE II and SOFA scores.Nevertheless, our study had several limitations. First, it was a single-center, retrospective, observational cohort study based in western China and thus our patient population may not be representative of TDL-MV patients in other regions or countries. Therefore, multi-center, prospective studies of TDL-MV patient prognostic risk factors are needed to externally validate the model. Second, the economic health of western China is relatively poor, especially in rural areas. As a result, some TDL patients resource-poor regions who need mechanical ventilation do not have access to ICUs and thus are under represented in ICU patient populations in such regions, thus leading tobias. Third, the prediction model was constructed using a retrospective design that would inevitably introduce bias into the results to thereby reduce the statistical strength of our conclusions. Fourth, although laboratory data may change as TB progresses, such changes are not adequately captured in retrospective study-based predictive models, since such studies cannot incorporate dynamic changes of indicators in the final analysis.

## CONCLUSION

In this study, a mortality prediction model based on independent risk factors of BMI < 18.5kg/m^2^, septic shock and a BUN level ≥7.14mmol/L was found to have good predictive power and differentiation ability. Moreover, internal validation of the model using retrospective data of adult TDL-MV patients admitted to the ICU of Chengdu Public Health Clinical Medical Center revealed that results obtained using the model were highly stable, reliable and reproducible. Thus, this model may be clinically useful for achieving early and rapid screening of patients with TDL who need mechanical ventilation and thus are at high risk of death.

## ACKNOWLEDGMENTS

We would like to thank the general practices and patients that participated in follow-up.

## FUNDING

This work was supported in part by special funds of Chengdu Science and Technology Bureau (2022-YF05-02139-SN), Chengdu Health Commission (2022101) and Sichuan Provincial Science and Technology Department (23ZDYF1334).

## AUTHOR CONTRIBUTION

KC, LJ and YZ received ethical approval for the study. KC, LY and YY participated in patient recruitment and data analysis. KC wrote the first draft of the manuscript. All authors reviewed and edited the manuscript and approved the final version of the manuscript.

## INSTITUTIONAL REVIEW BOARD STATEMENT

The study was carried out in accordance with the principles of the Declaration of Helsinki. This study was approved by the Medical Ethics Committee of Chengdu Public Health Clinical Medical Center (YJ-K2022-02-01).

## INFORMED CONSENT

Not applicable.

## DATA AVAILABILITY STATEMENT

The original contributions presented in the study are included in the article, and further inquiries may be directed to the appropriate authors.

## Notes

### Competing Interest Statement

The authors have declared no competing interest.

### Funding Statement

The funders had no role in study design, data collection and analysis, decision to publish, or preparation of the manuscript.

### Author Declarations

Ethics Committee of Chengdu Public Health Clinical Medical Center

